# Parameter Estimation for a Modified SEIR Model of the COVID-19 Dynamics in the Philippines using Genetic Algorithm

**DOI:** 10.1101/2022.05.17.22275187

**Authors:** Gabriel Lorenzo I. Santos

## Abstract

The Philippines has been under a series of different levels of community quarantine and this affected the dynamics of the COVID-19 spread in the country. Predicting the trajectory has been an interest of various research groups. To provide a more efficient method to estimate the parameters of the Age-Stratified, Quarantine-modified SEIR model with Nonlinear Incidence Rates (ASQ-SEIR-NLIR) other than the shooting method, a genetic algorithm approach is explored. By defining constraints for each parameter, the algorithm arrived at an acceptable optimal value for each parameter. The experiment is done on two regions of interest: the Philippines (country-level) and Quezon City, Metro Manila (city-level). The ASQ-SEIR-NLIR model, using the parameters generated by the genetic algorithm, is able to produce an average trajectory compared to the actual data, which may be deemed noisy. The dynamics of the COVID-19 spread between Quezon City and average country level is compared, showing that the city population is being exposed to the virus at a much faster rate than the country average and may have more asymptomatics not getting tested than the country average. Given the average trajectory, the peak daily infection projection is way lower at 0.0823% of the country population for the country projection and 0.1494% of the Quezon City population for the city projection, which is below than previous literature estimates of 3-10%.

## 1. INTRODUCTION

The first COVID case in the Philippines was detected on 30 January 2020 and the National Capital Region was placed under the Enhanced Community Quarantine (ECQ) on 15 March 2020, which was extended to the entire Luzon island on 16 March 2020. Since then, several research groups based in the Philippines have attempted to predict the rise of cases using various methods [1], and the most common ones are the time-series analysis [2, 3], compartmental models [4, 5, 6, 7, 8], and agent-based models [9, 10, 11].

### 1.1 The ASQ-SEIR-NLIR Model

#### 1.1.1 Overview

The Age-Stratified, Quarantine-modified SEIR model with Nonlinear Incidence Rates (ASQ-SEIR-NLIR) (see Figure 1) is a modification of the traditional Susceptible-Exposed-Infectious-Removed compartmental model to account for time-varying quarantine effect (*Q*(*t*)) [6], age-stratified infection probability (*U*) [7], and nonlinear incidence rates for behavioral (*α*) and disease-resistance (*ϵ*) factors [8]. These additional parameters serve as modification to the typical transmission rate (*β*), incubation rate (*σ*), and removal rate (*γ*) from the original SEIR model to provide a more realistic bounds for projections by incorporating impacts of real-world scenarios such as the ECQ and implementation of minimum health standards (MHS).

**Figure 1:**
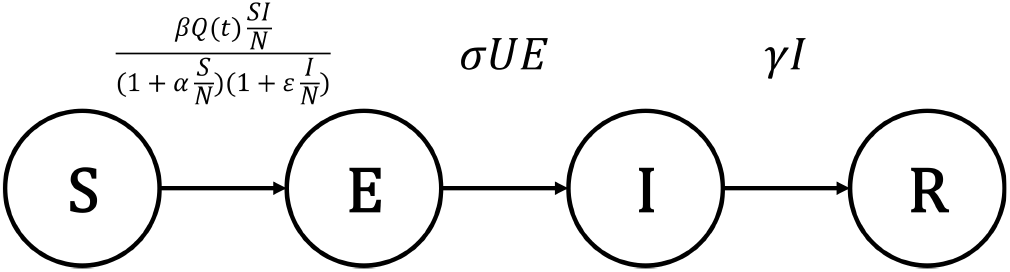
The ASQ-SEIR-NLIR Compartmental Model.

**Figure 2:**
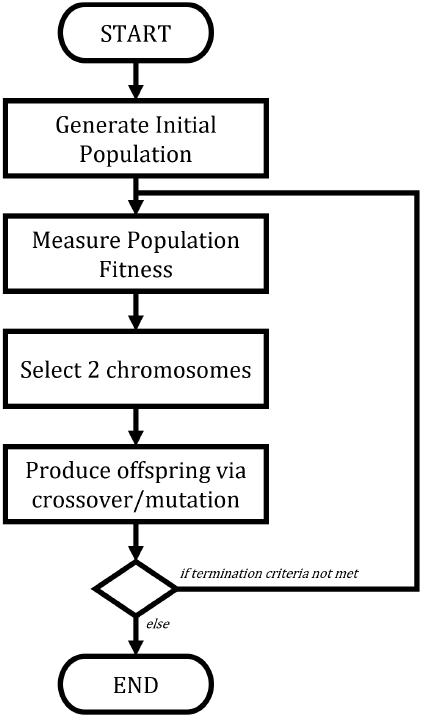
Five Phases of Genetic Algorithm.

#### 1.1.2 Governing ODEs

Given the transition rates between compartments, Equations 1-4 shows the governing ordinary differential equations of ASQ-SEIR-NLIR model.

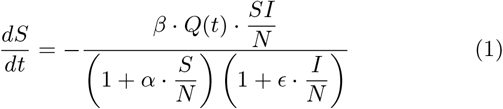

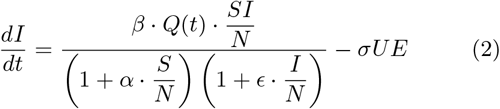

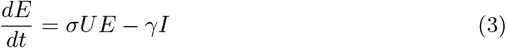

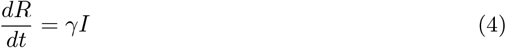

#### 1.1.3 Model Assumptions

This model assumes the following:

- The population is fixed and no birth and deaths are considered for the susceptibles. (N = S + E + I + R).
- Each compartment of SEIR are well-mixed and interact homogeneously.
- Infected individuals are removed by recovery with permanent immunity or by death
- The transmission rate (*β*), incubation rate (*σ*), and removal rate (*γ*) are inherent properties of the virus.

### 1.2 Genetic Algorithm

Genetic algorithm is a heuristic search or optimization technique that belongs to a larger class of evolutionary algorithms and relies on randomness in crossover and mutation processes between the highest quality or “fittest” chromosomes. It employs the Darwinian concept of natural evolution which involves five (5) processes: an initial population, a fitness (or objective) function, selection, crossover, and mutation [12]. Through these processes, the genetic algorithm makes new generation of chromosomes by making randomized changes to certain genes in the chromosomes belonging to the prior generation.

#### 1.2.1 Definitions

A *gene* is a single piece of data or a value of a single parameter. A *chromosome* (or individual), denoted by Ω, is a set of genes and refers to the set of values of all parameters explored in a specific environment or problem. A *population* is a set of chromosomes or individuals.

The genetic algorithm manipulates the chromosomes in a population by recombination or crossover and mutation of the resulting chromosomes. While it is common for genes to be represented by binary values, they can be represented by real continuous values, similar to the values of our parameters in the ASQ-SEIR-NLIR model.

### 1.3 Parameter Estimation

Ordinary differential equations, such as ASQ-SEIR-NLIR model, are treated as initial value problems and can be solved numerically using various forward approaches such as Euler’s method or Runge-Kutta methods. The output defines how the model behaves given a set of parameters and initial values; hence, they are called initial value problems. However, initial value problems do not use actual data, leaving the solution almost theoretical or bounded by wide ranges.

Ground truthing is necessary for the model behavior to resonate the reality to some degree. Ground truthing requires an objective. In this experiment, the model output has to be close to the actual data; therefore, the distance between the two should be as minimum as possible. In the context of the differential equation, ground truthing means achieving the set of parameters that will produce a trajectory close to the actual trajectory.

This ground truthing process may also be called parameter estimation. There are various ways to estimate parameters. One common technique is the shooting method, where the parameters are iteratively changed manually until an acceptable trajectory is achieved. As the definition says, this process may consume too much time of the analyst or computing resources if the optimal set of parameters are not achieved in a reasonable amount of time.

In this experiment, genetic algorithm is proposed to achieve the optimal set of parameters at a much more reasonable amount of time. Genetic algorithm is said to be a good alternative to gradient-based methods because of the stochasticity of genetic algorithms, giving genetic algorithms higher chances in finding a global optima [12]. A few things to note when implementing genetic algorithms are premature convergence, when the algorithm settles to early before reaching the global optima, and expensive computation due to multiple iterations [12].

## 2. METHODOLOGY

To implement the genetic algorithm and solve the differential equations, Python is primarily used, supported by Python libraries, specifically `numpy` and `scipy`. The experiment is conducted using a machine with AMD Ryzen 7 3700x chip at 3.6 GHz and 32 GB RAM at 3.6 GHz.

### 2.1 Data Preparation

The daily count for active cases, recovered, and deceased were extracted from the Department of Health COVID-19 Data Drop [14] using the tool developed by [9]. The daily active cases were transformed to the corresponding 7-day average to reduce the noise level of the data due to the case verification process of the government unit. The recovered and deceased count were added to form the removed count and was further transformed into daily cumulative removed cases. The dates extracted were from 07 March 2020 to 05 January 2022. The data were extracted last 07 January 2022, which means that the count prior case verification on that final week were only considered in this experiment and future changes due to the government agency’s verification process are disregarded.

### 2.2 Implementing the Genetic Algorithm

This section describes the implementation of the phases of genetic algorithm in the context of this experiment.

#### 2.2.1 Chromosome Representation

Given that there are seven (7) parameters in the ASQ-SEIR-NLIR model, the chromosome in this experiment consists of 7 floating-point-valued genes. Each gene is constrained with an acceptable ranges for each parameter of the ASQ-SEIR-NLIR model. Figure 3 shows a sample chromosome that shows the order of parameters and may be a potential solution to the problem.

**Figure 3:**
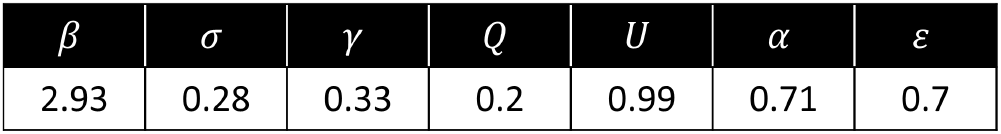
Chromosome with Sample Values.

One limitation of this representation is that it cannot represent any time-varying variable such as the quarantine score by [6]. It may be addressed by adding more genes representing a specific time interval such as 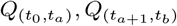 [12] but this modification is not pursued for this experiment because of the limitation of `scipy.integrate.odeint` solver not accepting array of values as parameter. Hence, the quarantine score *Q* produced by the genetic algorithm may be treated as an average for all data points, neglecting specific adjustments such as ECQ to Modified ECQ to General ECQ, etc.

#### 2.2.2 Fitness Function

An objective function in an optimization problem is a function where the maxima or minima is being searched within a certain domain. In genetic algorithm, this is called a fitness function, which generates the fitness score or quality score for each individual/chromosome. The fitness score is then used to determine which chromosomes are up for recombination or reproduction [12].

The fitness or quality of each chromosome is defined by comparing the results of the ASQ-SEIR-NLIR model on certain parameters with the actual data. This allows the algorithm to see how well would the ASQ-SEIR-NLIR model fits the reported data [13]. In this experiment, the actual data used as a comparison are the cumulative daily cases, *c*_*r*_, and cumulative removed (recovered and died, together), *r*_*r*_, cases. Their distance from the cumulative Infectious count, 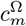, and recovered count, 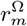, respectively, are measured using their *L*2 distance. This leaves the problem having a multi-objective function by having two sets of data being optimized. To get a single score for both fitness, a weighted sum of the two distances is calculated and optimized (see Equation 5).

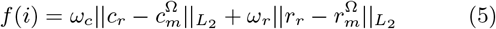

The fitness function *f* (*i*) [12] takes the distance between the predicted and actual cumulative active and removed cases and take their weighted sums. In this experiment, *ω*_*c*_, *ω*_*r*_ = 0.5 is used. To compute for 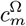 and 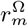, the chromosomes in the population Ω is used as input to the ASQ-SEIR-NLIR model. The outputs *I* (cumulative) and *R* is used as 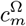 and 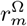, respectively.

The objective is to minimize the distance between the reported and predicted cases. Therefore, the fitness rank of a chromosome should be higher when its fitness score *f* (*i*) is smaller. Usually, the rank is assigned as an integer from rank 0 to *n*, where *n* is the population count and rank 0 is the highest. In this experiment, the rank is calculated as 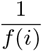. This automatically transforms the fitness score *f* (*i*) and makes the chromosome comply to the optimization rule (lower distance = higher rank). For the purpose of this experiment, this defines the difference of the fitness score and fitness rank, whereas the fitness score is the distance between the predicted and actual cases while the fitness rank is the position of the chromosome within the population based on their fitness score.

#### 2.2.3 Initial Population

A population size of 70 is used for this experiment. At this phase, each Ω = {*β, σ, γ, Q, U, α, ϵ*} is generated from a random sample of uniform distribution of values between the upper and lower bounds for each gene. The chosen ranges and their sources are shown on the table below.

| List of parameters and their ranges |  |  |
| --- | --- | --- |
| Parameter | Range | Source |
| Transmission/Contact Rate ($\beta$) | [2, 3.2] | [7, 8] |
| Incubation Period ($\sigma$) | [0.13, 0.28] | [5] |
| Removal Rate ($\gamma$) | [0.05, 0.33] | [8, 15] |
| Quarantine Factor ($Q$) | [0.2, 0.9] * | [7, 8] |
| Age-Stratified Infection Probability ($U$) | [0, 1] | [7] |
| Behavioral Factor ($\alpha$) | [0, 1] | [8] |
| Disease-resistance Factor ($\epsilon$) | [0, 0.7] * | [8] |

\* - author's preference, in addition to literature review

The range for *Q* is supposedly within [0, 1]. However, throughout the duration of the data, the country started with a very strict mobility restriction (estimated at *Q* = 0.2, which translates to 80% success [7, 8]) and has changed depends on the peak of cases and result of various interventions.

Behavioral factors refer to the use of face masks, compliance to social distancing, and maintaining hygiene [8]. It is also a reflection of various personal protective behaviors or compliance to minimum health standards. Literature indicates a range of *α* = [0.1, 0.6] [5]; however, the original range of *α* = [0, 1] [8] is used in the experiment.

Disease-resistance factors refer to proper nutrition, exercise, maintaining good overall health, vaccination, and other behavior related to boosting natural immunity [11]. Certain buying behaviors are observed within the duration of pandemic such as increased share of online fitness courses subscription [16] and other health-related purchases [17] even before vaccination officially started. When it comes to vaccination, 51% of the country population have received at least 1 dosage of vaccine [18]. To accommodate for some unaccounted factors, a range of *ϵ* = [0, 0.7] is chosen. A full *ϵ* = [0, 1] [8] is not selected due to vaccine supply limiting the increase in total vaccinated individuals.

#### 2.2.4 Selection

As part of the genetic algorithm, the selection process involves the selection of a pair of chromosomes randomly from the population. To ensure that only the fittest chromosomes are selected, the top 5 chromosomes according to their fitness ranks are selected to produce the children chromosomes of the next generation, including the 5 parent chromosomes. Selection of parent chromosomes is done through the Roulette Wheel selection, where the each chromosome takes a portion of the wheel based on their fitness ranks, indicating their probabilities of being selected. The higher the rank, the bigger the share in the wheel, the higher chances of getting selected. The roulette wheel script by [19] is modified (shown below) and implemented for this experiment.

**Code 1:**
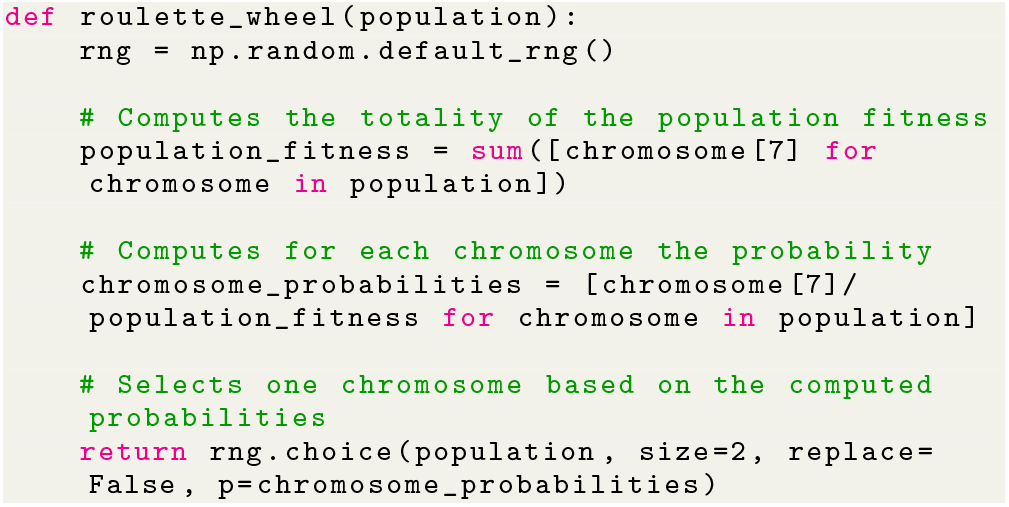
Roulette Wheel Implementation.

#### 2.2.5 Crossover

To produce the child chromosome, the parents has to be crossed over. On a simple genetic algorithm with chromosomes represented as binary strings, a typical crossover involves swapping of bits between the two parent chromosomes. For a floating point representation such as the representation used in this experiment, crossover or recombination involves taking a weighted sum of certain genes and replacing the original gene on each parent with the new recombined or crossed-over gene.

Not all parents encounter a crossover. First, a random floating-point valued number is generated. If the number meets the crossover probability threshold, only then the two parent chromosomes are crossed. The crossover is described by Equation 6 and 7. Given that *α* = 0.5 is used for this experiment, the crossover operator produces two (2) identical children chromosomes with genes calculated from the average of both parents’ genes. To reduce the chances of inserting duplicate chromosomes, only 1 child chromosome is sent for mutation.

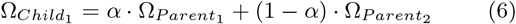

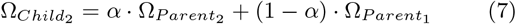

If the number previously generated does not meet the crossover probability, then the parent with higher fitness is forwarded for mutation.

#### 2.2.6 Mutation

Similar to the Darwinian concept of evolution, mutation may randomly exist during the reproduction of individuals or chromosomes. To implement this, the randomly-generated floating-point number is checked with the mutation probability threshold. It is meets the threshold, the child chromosome from the crossover operator is then set for mutation.

To mutate the chromosome, another random number is generated between 0 to 6 (representing the index of each gene in the chromosome). Then, each gene is assigned of a random number between 0 to 6. If the assigned number to a gene is greater than or equal to the random number previously generated, then these genes are adjusted to simulate mutation. The adjustment involves adding a number generated from a normal Gaussian distribution with spread or standard deviation equal to the mutation probability. If none of the assigned numbers to each gene meets the previously generated random number, then a random gene will be selected for mutation or adjustment.

If the mutation probability threshold is not met, then the resulting chromosome from the crossover process is then sent back to the population.

#### 2.2.7 Next Generation Population

As mentioned in Section 2.2.4, only the top 5 chromosomes from the prior generation are chosen for crossover and mutation. This means that to fill in the population size for the succeeding generation, the selection-crossover-mutation process is repeated until 65 children chromosomes are generated and completes the next generation population.

#### 2.2.8 GA Termination Criteria

The genetic algorithm is run continuously until either two (2) conditions occur: the algorithm has generated 2,500 generations, or the fitness score has surpassed the tolerance level. In this experiment, the tolerance level is 1e-6.

#### 2.2.9 Recursive Implementation of GA

After the initial population has been generated, the genetic algorithm runs fitness evaluation, selection, crossover, and mutation repeatedly through generations. For the sake of implementation simplicity and repeatability, a recursive implementation of GA is considered. This allowed the genetic algorithm to run using 1 line, making it easier to generate various scenarios given random selection and mutation.

The only limitation of using a recursive implementation is the high consumption of memory due to the stacking nature of recursive functions. This means that the data generated for all generations are stored until the break criteria is met. For this reason, the maximum generations is set to 2,500, instead of the previous value of 3,000.

#### 2.2.10 Experiment Runs

Due to the stochastic nature of selection and mutation process, it is possible that every run of the genetic algorithm produces different result. To address this, the experiment is run for five (5) times using the same initial population generated. This ensures that each run start with the same fitness function, while the randomness produces the different optimal parameters.

## 3. RESULTS AND DISCUSSIONS

Here, the results of the experiment is presented and the insights gathered are discussed. The experiment is run on two regions of interest: The Philippines (country level) and Quezon City, Metro Manila (municipality level).

### 3.1 Fitness Curves

The fitness curves show how the fitness score, defined as the distance between the actual data and predicted data, decreases after certain generations. Figure 4 shows how each run starts at the same fitness, indicating that each run uses the same initial population, as stated in Section 2.2.10, and continues stochastically until the termination criteria is met. Figure 4a shows that the most optimal fitness score among the 5 runs, using the country-level data, is achieved during the fifth run and that it has been converged between 400th to 500th generation. The fourth run is the last to converge and yet, has even come close to the objective function values the other 4 runs have achieved. Meanwhile, Figure 4b shows a different pattern, mainly due to the stochastic nature of selection and mutation. The most optimal run and earliest to converge is achieved by the 2nd run.

**Figure 4:**
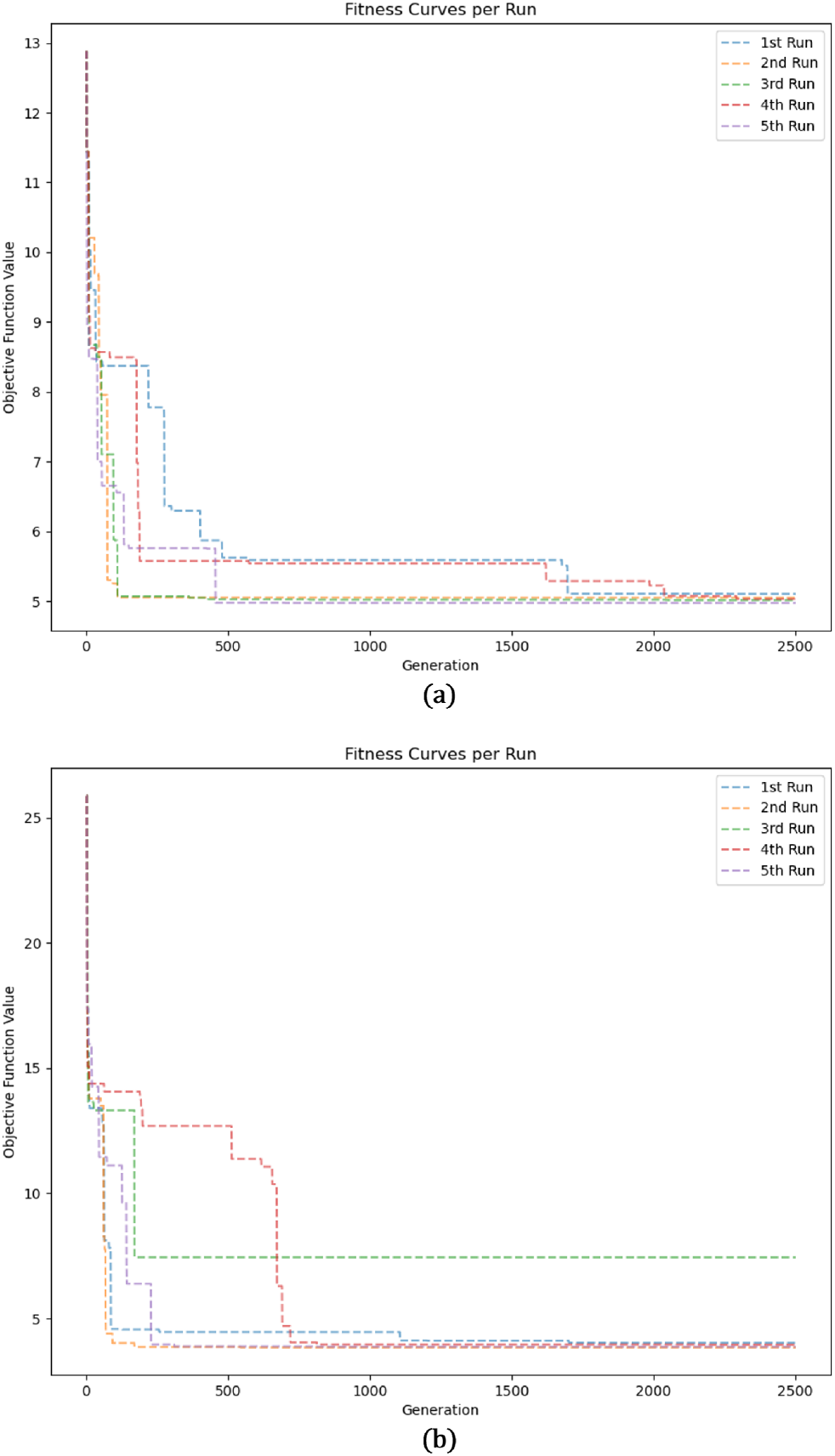
Fitness Curves for (a) The Philippines and (b) Quezon City.

### 3.2 ASQ-SEIR-NLIR Results

The optimal set of parameters is taken from the output of the most optimal runs, which are the 5th run for the country-level data, and 2nd run for the municipality-level data.

Figures 5 and 6 show the trajectory comparison between the actual data and predicted data from the ASQ-SEIR-NLIR model for the active cases and removed cases, respectively. The country-level fit is measured at 4.978. The municipality-level fit is measured at 3.857. The difference in fits may be traced back to the different peaks experienced by the Philippines, as a whole, and by Quezon City alone.

**Figure 5:**
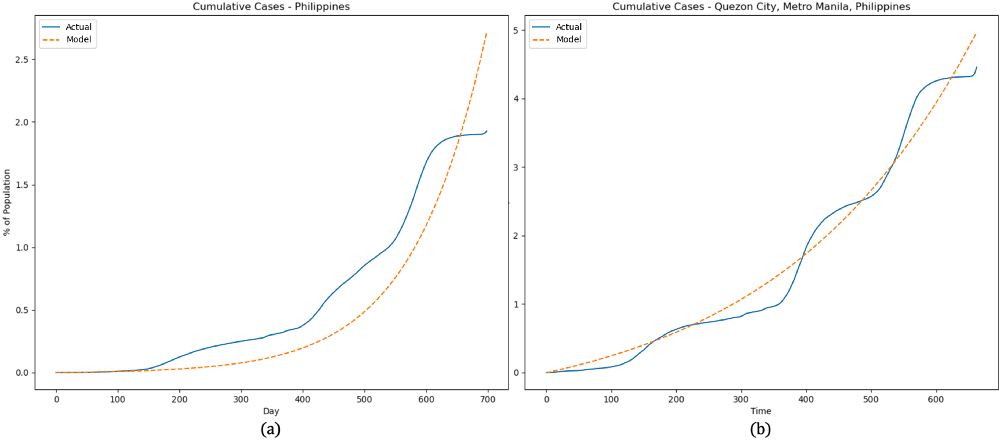
Cumulative Active Cases for (a) The Philippines and (b) Quezon City.

**Figure 6:**
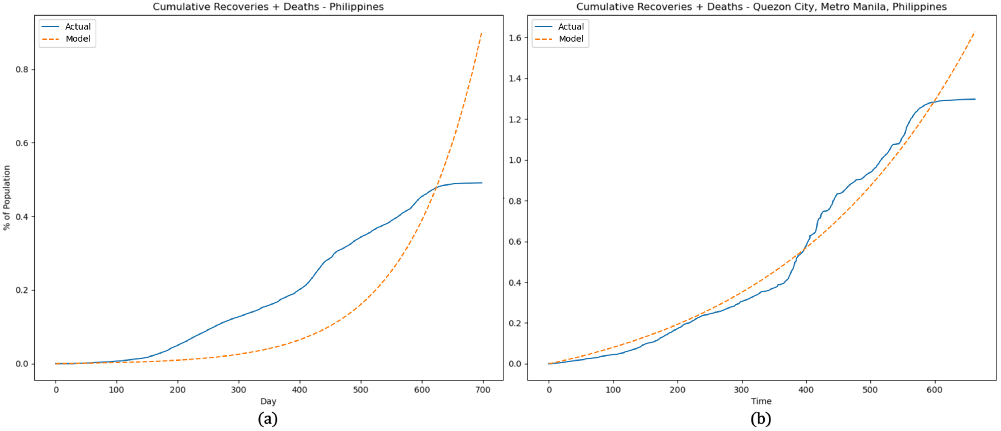
Cumulative Removed Cases for (a) The Philippines and (b) Quezon City.

As mentioned in Section 2.2.1, due to the limitation of `scipy.integrate.odeint` solver of not accepting a time-varying variable, an average trajectory is achieved. Figure 7 shows the actual daily cases against the predicted daily cases. It is noticeable how the trajectory produced by the ASQ-SEIR-NLIR model passes through the average of the actual data. This observation is valid for both country-level and citylevel.

**Figure 7:**
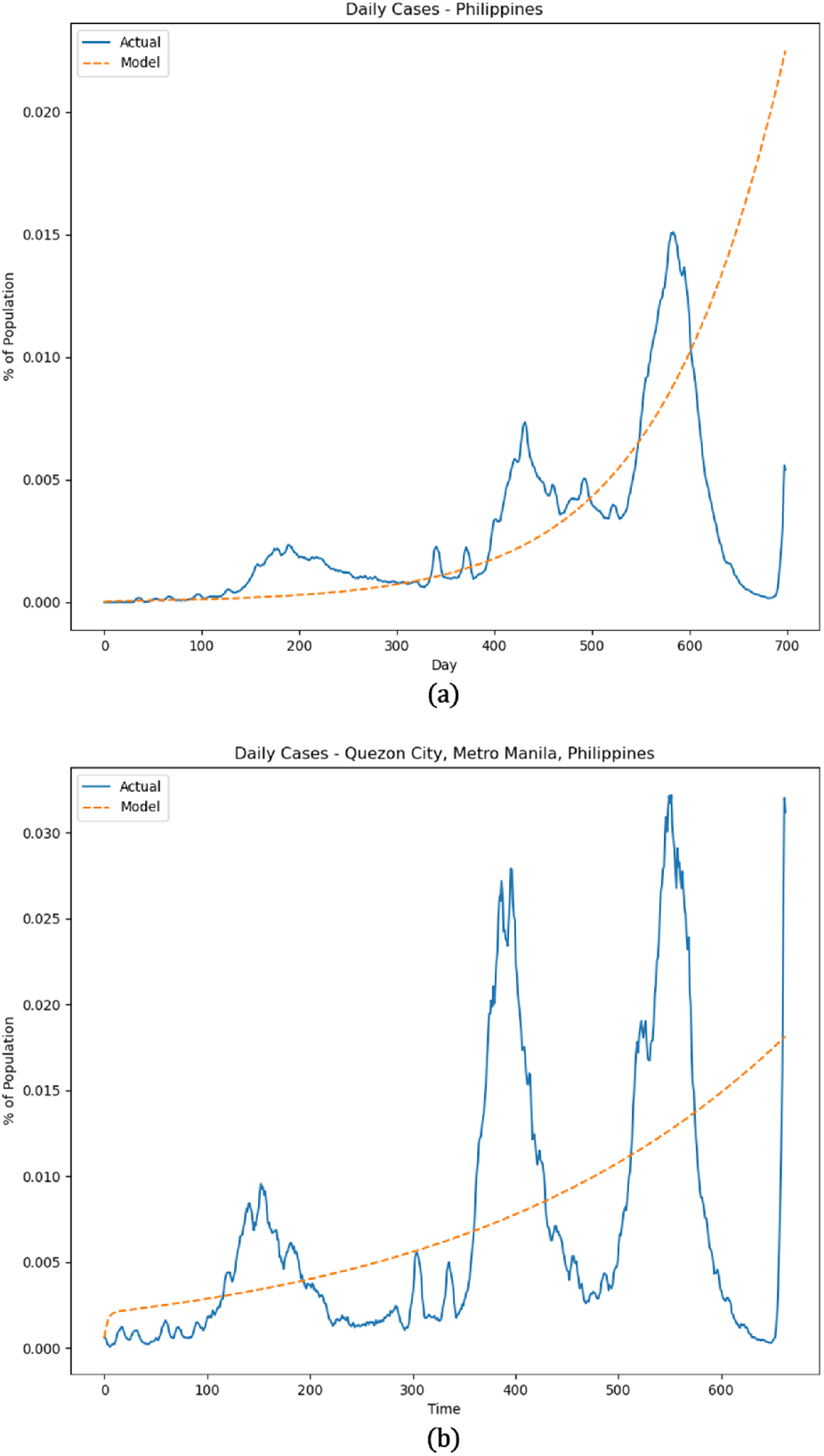
Daily Active Cases for (a) The Philippines and (b) Quezon City.

### 3.3 Dynamics of COVID-19 Spread

The optimal parameters obtained from the genetic algorithm help describe the dynamics of the spread of COVID-19 at the country-level and at city-level.

| Optimal Parameter Values |  |  |
| --- | --- | --- |
| Parameter | Country | City |
| Transmission/Contact Rate ($\beta$) | 2 | 3.12 |
| Incubation Period ($\sigma$) | 0.28 | 0.28 |
| Removal Rate ($\gamma$) | 0.33 | 0.33 |
| Quarantine Factor ($Q$) | 0.2 | 0.2 |
| Age-Stratified Infection Probability ($U$) | 1 | 0.03 |
| Behavioral Factor ($\alpha$) | 0.13 | 0.36 |
| Disease-resistance Factor ($\epsilon$) | 0.7 | 0.7 |

#### 3.3.1 Transition from Susceptible to Exposed

The parameters involved in this transition are the transmission or contact rate (*β*) and quarantine factor (*Q*), together with the nonlinear incidence rates behavioral factor (*α*) and disease-resistance factor (*ϵ*). Both regions of interest show relatively low compliance to minimum health standards yet relatively high disease-resistance rates, which may be attributed to increase vaccination efforts. This implies that there is high level of immunity but the high exposure rate is due to low compliance to minimum health standards. Both regions also differ in terms of contact rate but same quarantine factors. Quezon City has an average *β*_*QC*_ = 3.12, which means higher probability of getting exposed to the virus, versus the *β*_*PH*_ = 2 average for the whole country. However, the quarantine success spells the difference in terms of the total expected exposed individuals. Both Quezon City and the Philippines have an average of *Q*_*PH*_ = *Q*_*QC*_ = 0.2, indicating 80% quarantine success. Getting the product of the two parameters, the total transmission probability would be *βQ*_*PH*_ = 0.4 and *βQ*_*QC*_ = 0.624. This means that people in Quezon City are getting exposed to the virus faster compared to the country average.

#### 3.3.2 Transition from Exposed to Infectious

The parameters involved in this transition is the incubation period (*σ*) and age-stratified infection probability (*I*). Both the Philippines and Quezon City have an average 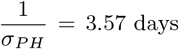. It is also observed that a city-level age-stratified infection probability could produce a lower range of values for the Infectious compartment than a country-level age-stratified infection probability. This is expected due to the averaging effect of aggregating granular data into the higher category, similar to aggregating city-level to country-level of information. Getting the product of the two parameters yields *σU*_*PH*_ = 0.28 and *σU*_*QC*_ = 0.0084. On average, more exposed individuals develop symptoms in the whole country than in Quezon City. However, it should be noted that these values are produced by the genetic algorithm from the confirmed detected cases. Asymptomatics who are not tested are not considered to have moved from Exposed to Infectious compartment. Given that assumption, it may be inferred that there may be more asymptomatic cases in Quezon City than in the Philippines, on average. However, this insight needs to be verified through other means.

#### 3.3.3 Transition from Infectious to Removed

The only parameter involved in this transition is the removal rate (*γ*). It shows that both Philippines and Quezon City has similar removal rate at *γ*_*PH*_ = *γ*_*QC*_ = 0.33. This could mean that the city is as capable of supporting their infected population as the whole country could support the whole nation. Since recovered and deaths compartments are combined, there is no way to identify which region is much more capable of bringing their infected population back to healthy state or the other way.

### 3.4 Peak Projections

Figure 8 shows the projections when the Infectious compartment peaks and at what level.

**Figure 8:**
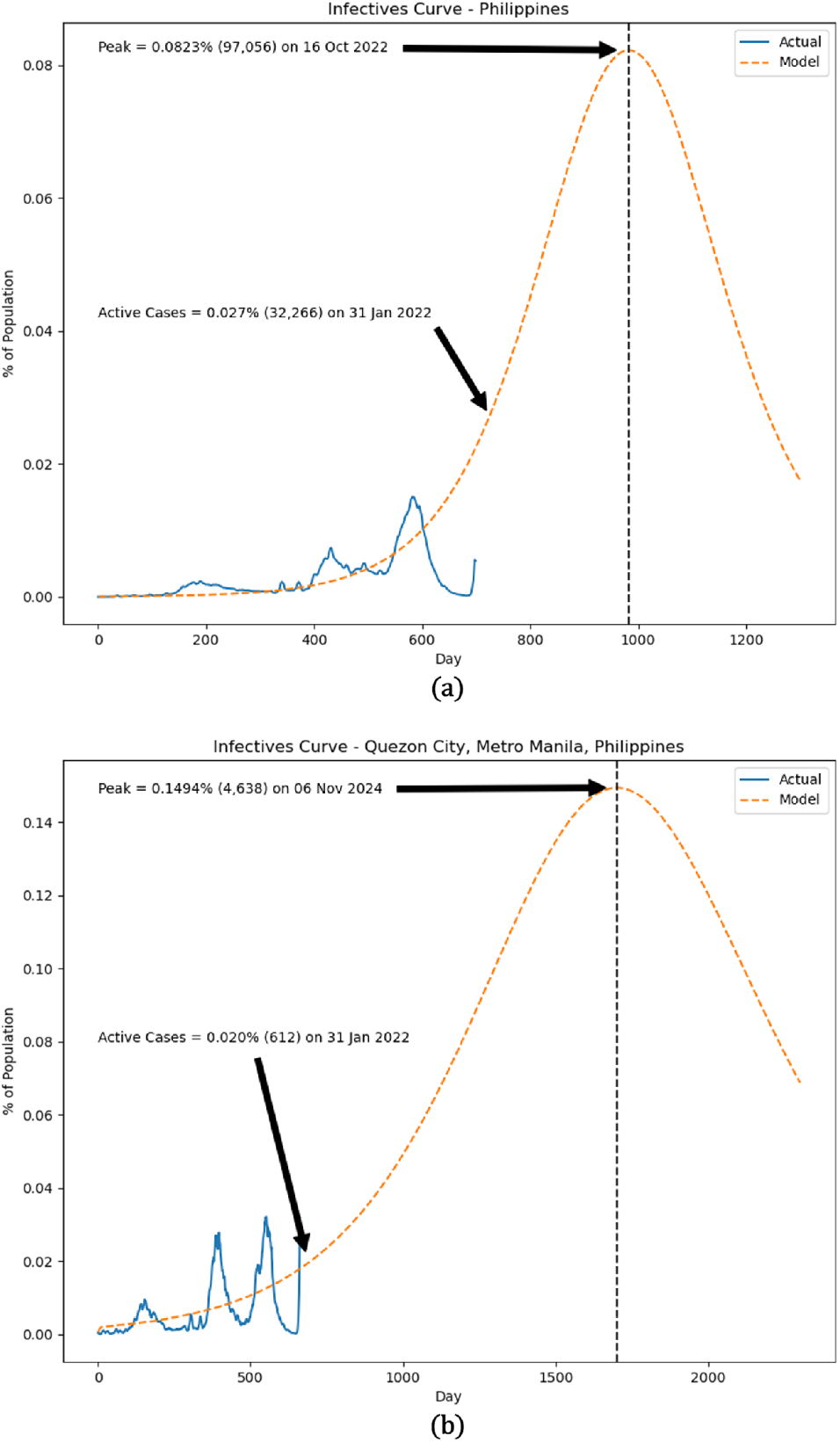
Peak Infection Projection for (a) The Philippines and (b) Quezon City.

It is observed that using the optimal parameters generated by the genetic algorithm, the country may experience a peak of average daily cases in October 2022 at 97,056 (0.0823% of the population). The peak cases is expected to grow to 30,000 by the end of January 2022, according to Jomar Rabajante, Ph.D of the University of the Philippines COVID-19 Pandemic Response team [20], which is echoed by the trajectory generated in this experiment at 32,266 (0.027% of the population).

Quezon City, on the other hand, may experience a peak of daily cases on November 2024 at 4,638 (0.1494% of the population). The Quezon City projection driven by parameters generated by the genetic algorithm is lower than previous literature projections of less than 10% [6], 5.791% [7], and 3.475% [8], indicating that the parameters generated by the genetic algorithm has produced a much flatter curve. It is also important to note that the previous projections [6, 7, 8] are generated using early 2020 data when the country is heavily reliant on community quarantine implementations and no variants of the original strain is present in the country. The lower projections can be attributed to the average impact of community quarantine, adherence to minimum health standards, and vaccination drive of each region of interest.

## 4. SUMMARY AND FURTHER WORK

In this experiment, a genetic algorithm approach is used to estimate the parameters of the ASQ-SEIR-NLIR model based on actual data provided by the Philippines’ Department of Health. While the parameters fails to fit the data on individual peaks, it is able to capture the average dynamics of the COVID-19 spread in the Philippines and within Quezon City, Metro Manila, given the difference in community quarantine implementations, adherence to minimum health standards, and vaccination completion rates. The parameters generated by the genetic algorithm also provide insights on the dynamics of the COVID-19 spread at country-level and at city-level. First, it is identified that the population in Quezon City is exposed to the virus at a much faster rate than the average exposure time of the country. It is also observed that while Quezon City and the whole country has an incubation period of 3.57 days, the resulting transition rate is much lower in Quezon City than the country average, which may be attributed to more asymptomatics in Quezon City not getting tested. Both Quezon City and the overall country has a removal rate of 0.33, showing the capability of the municipality to support infected population at the same level as the rest of the country. Lastly, the model generates projections for the country similar to other current projections, and lower peak projection for Quezon City, which may be interpreted as a better outcome than what the previous literature has expected.

To further improve the results, the trajectory of the model has to fit somehow with the individual peaks. This is possible if time-specific parameters are considered and a specific solver is developed to incorporate time-varying inputs. Other fitness function may be explored to acknowledge the nonlinearity of the dynamics and the uncertainty in actual data due to unreported cases and inefficient contact tracing. Lastly, the ASQ-SEIR-NLIR model itself can be modified further to capture other compartments such as asymptomatics, vaccinated, recovered, and deaths.

## Data Availability

All data produced in the present study are available upon reasonable request to the authors.

## REFERENCES

[1] Monica C. Torres, Christian Alvin H. Buhat, Ben Paul B. Dela Cruz, Edd Francis O. Felix, Eleanor B. Gemida, and Jonathan B. Mamplata. 2020. Forecasting COVID-19 cases in the Philippines using various mathematical models. medRxiv (2020). DOI:http://dx.doi.org/10.1101/2020.10.07.20208421

[2] J. Mikhail Solitario. 2020. Modified Community Quarantine beyond April 30: Analysis and Recommendations. (April 2020). Retrieved Janury 4, 2022 from https://up.edu.ph/modified-community-quarantinebeyond-april-30-analysis-and-recommendations/

[3] Parikshit Gautam Jamdade and Shrinivas Gautamrao Jamdade. 2021. Modeling and prediction of COVID-19 spread in the Philippines by October 13, 2020, by using the VARMAX time series method with preventive measures. Results in Physics 20 (2021), 103694. DOI:http://dx.doi.org/10.1016/j.rinp.2020.103694

[4] J. Arcede, et al. 2020. Accounting for symptomatic and asymptomatic in a SEIR-type model of COVID-19. Mathematical Modelling of Natural Phenomena. 15, 34, DOI:https://doi.org/10.1051/mmnp/2020021

[5] J. Caldwell, et al. 2021. Understanding COVID-19 dynamics and the effects of interventions in the Philippines: A mathematical modelling study. The Lancet Regional Health - Western Pacific. 14, 100211, DOI:https://doi.org/10.1016/j.lanwpc.2021.100211

[6] V. Bongolan, et al. 2021. Age-Stratified Infection Probabilities Combined With a Quarantine-Modified Model for COVID-19 Needs Assessments: Model Development Study. Journal of Medical Internet Research. 23, 5 (2021), e19544. DOI:http://doi.org/10.2196/19544

[7] J. Rayo, et al. 2020. Modeling the dynamics of COVID-19 using Q-SEIR model with age-stratified infection probability. medRxiv. DOI:http://doi.org/10.1101/2020.05.20.20095406

[8] J. Minoza, et al. 2020. Protection after Quarantine: Insights from a Q-SEIR Model with Nonlinear Incidence Rates Applied to COVID-19. medRxiv. DOI:http://doi.org/10.1101/2020.06.06.2012

[9] J. Celeste and V. Bongolan. 2021. School Re-opening Simulations with COVID-19 Agent-based Model for Quezon City, Philippines. The International Archives of the Photogrammetry, Remote Sensing and Spatial Information Sciences. XLVI-4/W6-2021, (2021), 85-90. DOI:https://doi.org/10.5194/isprs-archives-XLVI-4-W6-2021-85-2021

[10] V.P. Bongolan et al. 2021. COVID-19 AGENT-BASED MODEL: AN EPIDEMIOLOGICAL SIMULATOR APPLIED IN VACCINATION SCENARIOS FOR QUEZON CITY, PHILIPPINES. (November 2021). Retrieved January 4, 2022 from https://doi.org/10.5194/isprs-archives-XLVI-4-W6-2021-65-2021

[11] J. Minoza, et al. 2020. COVID-19 Agent-Based Model with Multi-objective Optimization for Vaccine Distribution. (April 2020). Retrieved January 4, 2022 from. https://www.researchgate.net/publication/348832487_COVID-19_Agent-Based_Model_with_Multi-objective_Optimization_for_Vaccine_Distribution

[12] D. Spataru. 2021. Using a Genetic Algorithm for Parameter Estimation in a Modified SEIR Model of COVID-19 Spread in Ontario. University of Guelph.

[13] P. Yarsky. 2021. Using a genetic algorithm to fit parameters of a COVID-19 SEIR model for US states. Mathematics and Computers in Simulation 185 (2021), 687–695. DOI:http://dx.doi.org/10.1016/j.matcom.2021.01.022

[14] Department of Health. COVID-19 Tracker: Department of Health website. Retrieved January 4, 2022 from https://doh.gov.ph/covid19tracker

[15] N.J.L. Haw, J. Uy, K.T.L. Sy, and M.R.M. Abrigo. 2020. Epidemiological profile and transmission dynamics of COVID-19 in the Philippines. Epidemiology and Infection 148 (September 2020), e204. DOI:http://dx.doi.org/10.1017/s0950268820002137

[16] Statista. 2021. Philippines: share of people using online gym courses during COVID-19 2020. (September 2021). Retrieved January 5, 2022 from https://www.statista.com/statistics/1186438/philippines-share-of-people-using-online-gym-courses-covid-19/

[17] Manila Bulletin. 2020. Shopee identifies 4 of the biggest e-commerce trends in 2020. Manila Bulletin (May 2020).

[18] Mathieu, E., Ritchie, H., Ortiz-Ospina, E. et al. 2021. A global database of COVID-19 vaccinations. Nat Hum Behav (2021)

[19] Roc Reguant. 2021. Roulette Wheel Selection in Python. (March 2021). Retrieved January 5, 2022 from https://rocreguant.com/roulette-wheel-selection-python/2019/

[20] Xave Gregorio. 2022. DOH projects COVID-19 cases to peak end-January, may surpass Delta spike. Philippine Star (January 2022).

